# Epigenetic Clock CpGs form Tumor methylation Programs that Predict Survival Across Cancers

**DOI:** 10.64898/2026.06.01.26354667

**Authors:** Pranava Gande, Alfred Kao, Omar Mokhashi, Wei Tse Li, Weg M. Ongkeko

## Abstract

Epigenetic clocks have been widely evaluated as cancer biomarkers, but findings have been inconsistent across tumor types and clinical endpoints. This inconsistency may reflect a fundamental misspecification: standard clock analyses treat clock CpGs as scalar aging readouts, assuming that tumors shift along the same methylation-aging axis as normal tissue. We tested this assumption across nine TCGA (The Cancer Genome Atlas) cancer types using Horvath clock CpGs. Tumors did not show a consistent mean shift in Horvath age acceleration relative to normal tissue. Instead, they showed an order-of-magnitude increase in age-acceleration variance. Scalar clock summaries also failed as survival biomarkers, producing zero nominal associations after adjustment for age, sex, and stage.

We therefore analyzed Horvath clock CpGs as a coordinated methylation system. PCA-derived tumor methylation programs captured 34.9%–50.8% of clock-CpG variance across cancers, and this structure persisted after adjustment for tumor purity, proliferative history, and global methylation instability. This indicates that tumor-associated clock-CpG variation is not random methylation disorder, but is organized along major axes of covariation. In fully adjusted Cox models, tumor methylation programs produced 15 nominal survival associations, four of which remained significant after FDR correction. Survival-associated programs were linked to transcriptional pathway activity, including UPR, EMT, IFN-γ response, and stemness, in directions consistent with their survival effects. These programs also persisted after controlling for tumor-normal differentially methylated regions, indicating that they were not explained by average cancer-associated methylation shifts. External validation in GSE72308 showed that two of three TCGA-nominated BRCA survival associations reproduced in the same protective direction.

The dominance of tumor methylation programs over scalar clock summaries shows that the cancer-relevant signal in epigenetic clock CpGs is not methylation age, but coordinated tumor-state structure. In tumors, Horvath’s clock CpGs are reorganized into methylation programs that capture survival-relevant and transcriptionally linked cancer biology.

## Introduction

DNA methylation is widely used for cancer biomarker development because tumor methylation patterns are stable, preserve tissue-of-origin information, and can be measured from fragmented DNA, including circulating tumor DNA [1]. Liquid biopsy-based methylation profiling is increasingly preferred in clinical settings over solid tissue samples due to its non-invasive nature [2,3]. Epigenetic clocks are especially attractive in this setting because they provide predefined CpG sets with established behavior in normal tissues [4]. In cancer, however, clock-derived biomarkers have produced inconsistent results across tumor types and clinical endpoints [5-7].

Most cancer applications interpret epigenetic clocks through methylation age or age acceleration, asking whether tumors appear epigenetically older or younger than expected [8]. This framing assumes that tumor-associated changes in clock CpGs lie along the same methylation-aging axis observed in normal tissues. If that assumption fails, scalar clock summaries may discard the tumor-relevant information contained in the CpG set.

Tumors may alter clock CpGs through a different mode of disruption. Rather than shifting uniformly toward older or younger methylation age, cancer may break the normal coordination that allows clock CpGs to function as an age predictor and reorganize them into tumor-specific methylation structure. This is plausible given the widespread epigenetic remodeling, methylation instability, developmental reprogramming, and adaptive cell-state transitions that characterize tumors [9-11].

Horvath’s clock was the first epigenetic clock developed by Steve Horvath in 2013, consisting of 353 CpG sites that estimate pan-tissue biological age [12]. In this study, we tested whether Horvath’s clock CpGs contain coordinated tumor methylation programs linked to survival and transcriptional state across cancers. Using TCGA methylation data from nine cancer types, we first evaluated whether conventional scalar clock measurements predicted overall survival. We then defined tumor methylation programs by PCA of Horvath clock-CpG methylation variation within each cancer type and tested whether these programs captured structured methylation variation, predicted survival, associated with transcriptional pathway activity, persisted after tumor-normal DMR control, and reproduced in an independent BRCA cohort.

## Methods

### Data sources and sample preprocessing

Illumina HumanMethylation450 (450K) DNA methylation data for Bladder Urothelial Carcinoma (BLCA, normal = 21, tumor = 416), Lung Squamous Cell Carcinoma (LUSC, normal = 42, tumor = 370), Lung Adenocarcinoma (LUAD, normal = 32, tumor = 471), Breast Invasive Carcinoma (BRCA, normal = 97, tumor = 796), Liver Hepatocellular Carcinoma (LIHC, normal = 50, tumor = 380), Colon Adenocarcinoma (COAD, normal = 38, tumor = 308), Kidney Renal Clear Cell Carcinoma (KIRC, normal = 160, tumor = 323), Head and Neck Squamous Cell Carcinoma (HNSC, normal = 50, tumor = 530), and Skin Cutaneous Melanoma (SKCM, normal = 2, tumor = 473) were obtained from The Cancer Genome Atlas (TCGA) via the UCSC Xena Browser. The SKCM cohort was not used for tumor-normal comparisons due to insufficient sample size of normal tissue. Corresponding clinical metadata, including sample type (tumor vs. normal) and age at diagnosis, were retrieved from the Genomic Data Commons (GDC) Data Center.

Clock-CpG analyses used the subset of available probes in each cancer-type methylation matrix. DNA methylation data were represented as β-values for methylation clock calculations. For PCA-based tumor methylation program analysis, Horvath clock CpGs with >5% missingness or β-value SD ≤0.02 were removed, and any remaining missing values were median-imputed within cancer type. β-values were converted to M-values, batch-corrected within cancer type when batch variable was available, and centered and scaled across tumor samples before PCA.

### Epigenetic clock calculation and scalar clock summaries

Epigenetic age was estimated using Horvath’s multi-tissue DNA methylation age clock (Horvath1), computed from median-imputed β-values using the *methylCIPHER* package [13]. Age acceleration was defined as the residual from a linear regression of Horvath1 methylation age on chronological age, fit in normal samples and applied to both tumor and normal samples within each cancer type.

Scalar clock summaries were used as comparator biomarkers in survival analysis. These were Horvath1 methylation age, Horvath1 age acceleration, absolute Horvath1 age acceleration, mean Horvath clock-CpG methylation, and within-sample Horvath clock-CpG methylation variability. Mean methylation and methylation variability were calculated using β-values.

### Covariates and tumor-level confounding

Age, sex, and pathologic stage were used as clinical covariates. Tumor purity estimates were obtained from TCGAbiolinks, prioritizing ABSOLUTE estimates where available and using Consensus Purity Estimates (CPE) otherwise. Proliferative history was measured using epiTOC2, a DNA methylation clock that estimates cumulative mitotic divisions based on cell division-associated methylation patterns [14].

Global methylation instability was quantified as the mean absolute deviation of each sample’s methylation values from a normal reference. For each cancer type, normal-sample mean β-values were computed at each CpG, and each sample’s InstabIndex was calculated as the average absolute deviation from this reference across 20,000 randomly sampled CpGs after missingness filtering and median imputation.

### Tumor methylation program definition

Tumor methylation programs were defined as the principal components of Horvath clock-CpG methylation variation within each cancer type. PCA was performed on tumor samples using batch-corrected Horvath CpG M-values centered and scaled across tumors. Each program captures a coordinated pattern of clock-CpG methylation variation across tumors. The ten PCs explaining the most variance were used as sample-level program scores for downstream analyses. PCA was also performed after residualization Horvath CpG methylation values for tumor purity, epiTOC2, and InstabIndex to measure adjusted PC1-PC10 variance.

### Survival analysis

Overall survival associations were tested separately within each cancer type using Cox proportional hazards models. PC1-PC10 tumor methylation program scores from the primary PCA were tested individually. Hazard ratios are reported per one-standard-deviation increase in program score. Models were fit unadjusted, adjusted for age, sex, and pathologic stage, and fully adjusted for age, sex, stage, tumor purity, epiTOC2, and InstabIndex. Cox models used complete cases for overall survival time, vital status, and model covariates. P-values were adjusted using the Benjamini-Hochberg false discovery rate method. Fully adjusted models were used as the primary survival analysis. Representative Kaplan-Meier curves show survival stratification after classifying patients into high- and low-score groups by the median program score.

### Transcriptional signature analysis

Gene expression data for TCGA HNSC, LUAD, and COAD were obtained as STAR FPKM-UQ matrices, mapped to gene symbols, and log2(x+1)-transformed before GSVA (Gene Set Variation Analysis). GSVA was run separately within each cancer type for four predefined signatures: IFNγ response, unfolded protein response (UPR), epithelial–mesenchymal transition (EMT), and curated stemness markers. IFNγ, UPR, and EMT were taken from the MSigDB Hallmark collection [15], and stemness was defined using MALTA_CURATED_STEMNESS_MARKERS [16].

Associations between survival-associated tumor methylation programs and GSVA scores were tested in tumor samples using Spearman’s correlation and partial correlation adjusted for age, sex, pathologic stage, tumor purity, epiTOC2, and InstabIndex. P-values were adjusted across signatures within each program using the Benjamini-Hochberg false discovery rate method.

### DMR-control sensitivity analysis

To control for tumor-normal differentially methylation regions, tumor-normal mean differences were estimated at Horvath clock CpGs within each cancer type and removed before PCA for this sensitivity analysis. PCA was repeated on the DMR-adjusted matrix using the same tumor-only scaling as the primary analysis.

### External validation

External validation was performed in GSE72308, an independent BRCA methylation cohort with overall survival data (n=295, dead = 52, alive = 239, null = 4). TCGA-BRCA programs nominally associated with survival after controlling for age, sex, stage, tumor purity, proliferative history and global methylation instability were selected for validation. GSE72308 samples were restricted to overlapping Horvath CpGs, converted to M-values, scaled using TCGA-BRCA training parameters, and projected onto the original TCGA-BRCA PCA loadings. PCA was not refit in GSE72308. Projected scores were tested for overall survival.

### Statistical analysis and visualization

All analyses were performed in R version 4.5.2. Statistical tests were two-sided, and multiple testing was controlled using the Benjamini-Hochberg false discovery rate method.

Tumor-normal differences in age-acceleration variability were tested using Fligner-Killeen tests. Age-acceleration mean shifts were tested within each cancer type. Correlations were assessed using Spearman correlation or covariate-adjusted partial correlation. Figures were generated using *ggplot2*.

## Results

### Tumors break epigenetic clocks through dispersion, not age acceleration

Tumors showed a striking expansion in Horvath age acceleration variability compared to normal, corresponding to 5.3x to 23.5x higher variance across eight cancer types. The increased dispersion was highly significant in BLCA (SD 5.27→25.54; Fligner p=4.33×10^−6^), LUSC (3.60→15.86; p=2.28×10^−10^), LUAD (4.88→21.30; p=2.03×10^−7^), BRCA (5.56→20.66; p=1.80×10^−16^), LIHC (5.42→19.86; p=5.63×10^−9^), COAD (4.95→17.24; p=1.48×10^−8^), KIRC (4.41→13.15; p=1.25×10^−23^), and HNSC (7.85→18.00; p=1.99×10^−6^).

Mean age acceleration did not show a consistent shift across cancers. Using p<0.05 as the significance threshold, BLCA (0→−7.89; p=0.0123), LUSC (0→−16.41; p=1.38×10^−15^), COAD (0→−4.91; p=0.0142), and HNSC (0→−11.31; p=8.66×10^−8^) were significantly negatively shifted, LIHC (0→6.87; p=0.00252) and KIRC (0→11.24; p=2.82×10^−27^) were significantly positively shifted, and LUAD (p=0.986) and BRCA (p=0.169) were not significantly shifted.

**Fig. 1.**
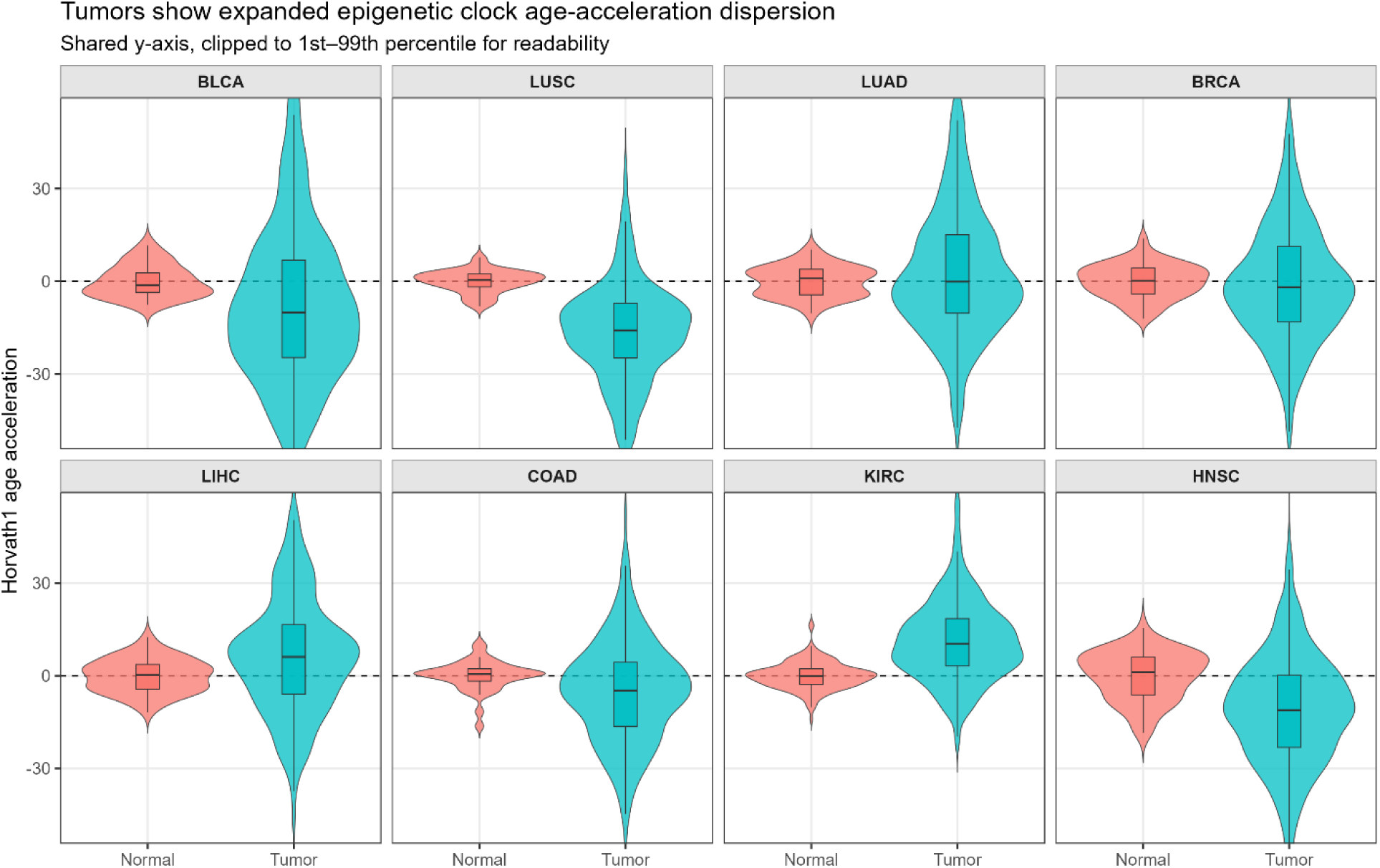
Tumors show expanded epigenetic clock age-acceleration dispersion. Horvath age-acceleration distributions show a marked expansion in variance in tumors (BLCA SD 5.27→25.54, LUSC 3.60→15.86, LUAD 4.88→21.30, BRCA 5.56→20.66, LIHC 5.42→19.86, COAD 4.95→17.24, KIRC 4.41→13.15, HNSC 7.85→18.00; all Fligner p<10^−5^). Mean shifts are inconsistent and absent in LUAD (p=0.986) and BRCA (p=0.169).

### Scalar clocks fail as survival biomarkers

Because tumors showed age-acceleration dispersion, absolute age acceleration was tested for prognostic relevance. Four other scalar clock measurements were also tested: clock age, age acceleration, mean methylation across clock CpGs, and within-sample clock-CpG methylation variability. Across nine cancers, unadjusted Cox models produced six nominal associations from 43 tests (p < 0.05), but none survived FDR correction. When adjusted for age, sex, and stage, none remained nominally significant.

**Fig 2.**
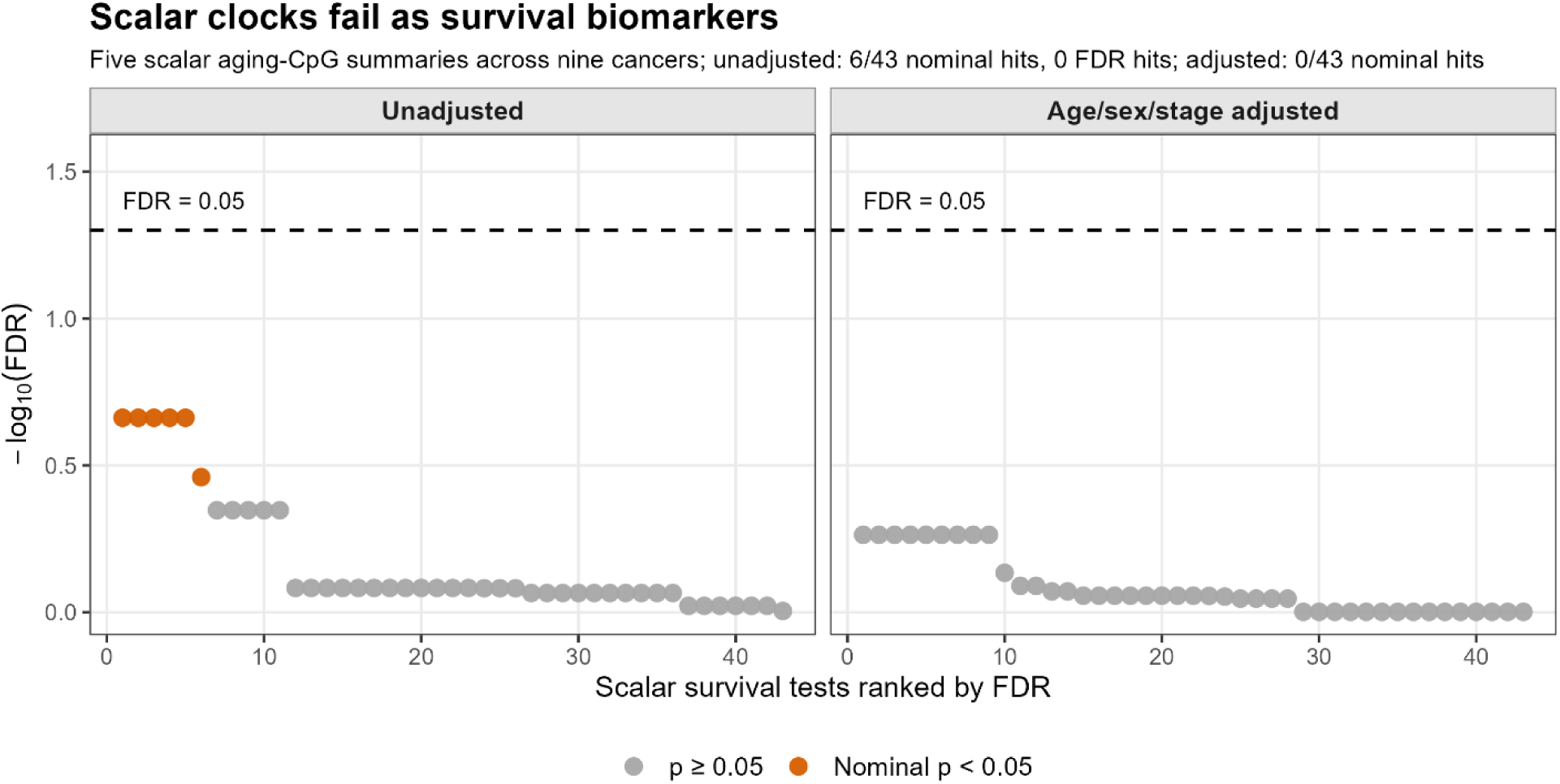
Scalar clocks fail as survival biomarkers. Ranked FDR values from Cox models testing five scalar aging-CpG summaries across nine cancers. Each point represents one test. Unadjusted and age/sex/stage-adjusted models are shown separately. Orange points indicate nominal associations (p < 0.05); gray points indicate non-nominal tests. In unadjusted models, six of 43 scalar tests were nominally significant, but none survived FDR correction. After adjustment for age, sex, and stage, no scalar clock summary remained nominally significant.

### Clock CpGs form tumor methylation programs

Epigenetic clock dispersion might arise from nonspecific methylation instability. PCA was performed within each cancer type on batch-corrected Horvath CpG values scaled across tumor samples, and these PCA-derived axes were defined as tumor methylation programs. Across nine cancers, PCA used 104-791 tumor samples and 271-292 Horvath clock CpGs per cancer. PC1 explained 9.7%-17.9% of tumor clock-CpG variance, and PC1-PC10 explained 34.9%-50.8%. After adjustment for tumor purity, proliferative history, and global methylation instability, PC1-PC10 still explained 28.2%-46.1% of variance. Thus, clock-CpG variation was not simply independent CpG-level noise or a byproduct of tumor purity, proliferative history, or global methylation instability, but instead formed coordinated tumor methylation programs.

**Fig 3.**
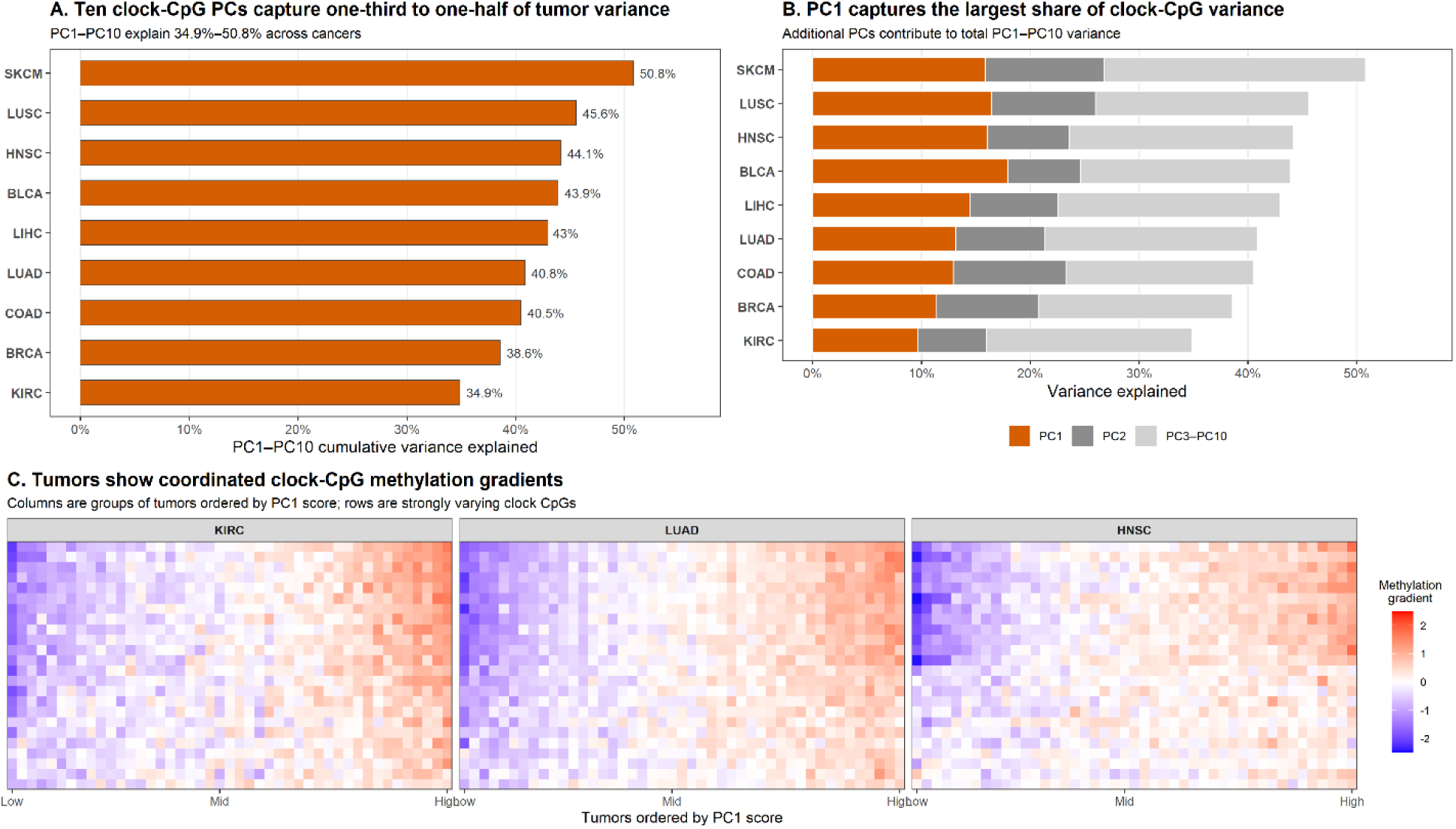
Clock CpGs form tumor methylation programs. (A) PC1-PC10 captured 34.9%-50.8% of clock-CpG variance across nine cancers. (B) PC1 explained the largest share of variance in each cancer, while additional PCs contributed to the total PC1-PC10 variance. (C) Ordering tumors by PC1 revealed smooth clock-CpG methylation gradients, supporting coordinated methylation structure rather than independent CpG-level variation. After adjustment for tumor purity, proliferative history, and global methylation instability, PC1-PC10 still explained 28.2%-46.1% of variance.

### Tumor methylation programs predict survival

Ten tumor methylation programs were analyzed in each of nine cancers using Cox models. After adjustment for age, sex, and stage, 13 associations were nominally significant and four survived global FDR correction. HNSC PC4 and KIRC PC1 were associated with worse survival (HNSC PC4: HR 1.35, 95% CI 1.16–1.57, p=9.93×10^−5^, FDR=0.011; KIRC PC1: HR 1.41, 95% CI 1.14–1.74, p=0.00137, FDR=0.038). LUAD PC4 and LUAD PC9 were associated with improved survival (LUAD PC4: HR 0.76, 95% CI 0.65–0.88, p=1.94×10^−4^, FDR=0.014; LUAD PC9: HR 0.75, 95% CI 0.64–0.88, p=3.30×10^−4^, FDR=0.019).

After further adjustment for tumor purity, proliferative history, and global methylation instability, 15 associations were nominally significant and four survived global FDR correction. In the full model, HNSC PC4 strongly predicted worse survival (HR 1.49, 95% CI 1.25–1.78; p=8.01×10^−6^; FDR=0.0028), while LUAD PC4, LUAD PC9, and COAD PC1 were associated with improved survival (HR 0.74, 0.76, and 0.60; all FDR≤0.027). Nominal associations were observed in eight of nine cancers. This contrasted with scalar clock measurements, which showed no nominal survival associations after clinical adjustment. Proliferative history and global methylation instability also failed as standalone biomarkers in clinically adjusted Cox models, producing no nominal associations across 17 tests (minimum p=0.126; minimum FDR=0.593).

**Fig. 4.**
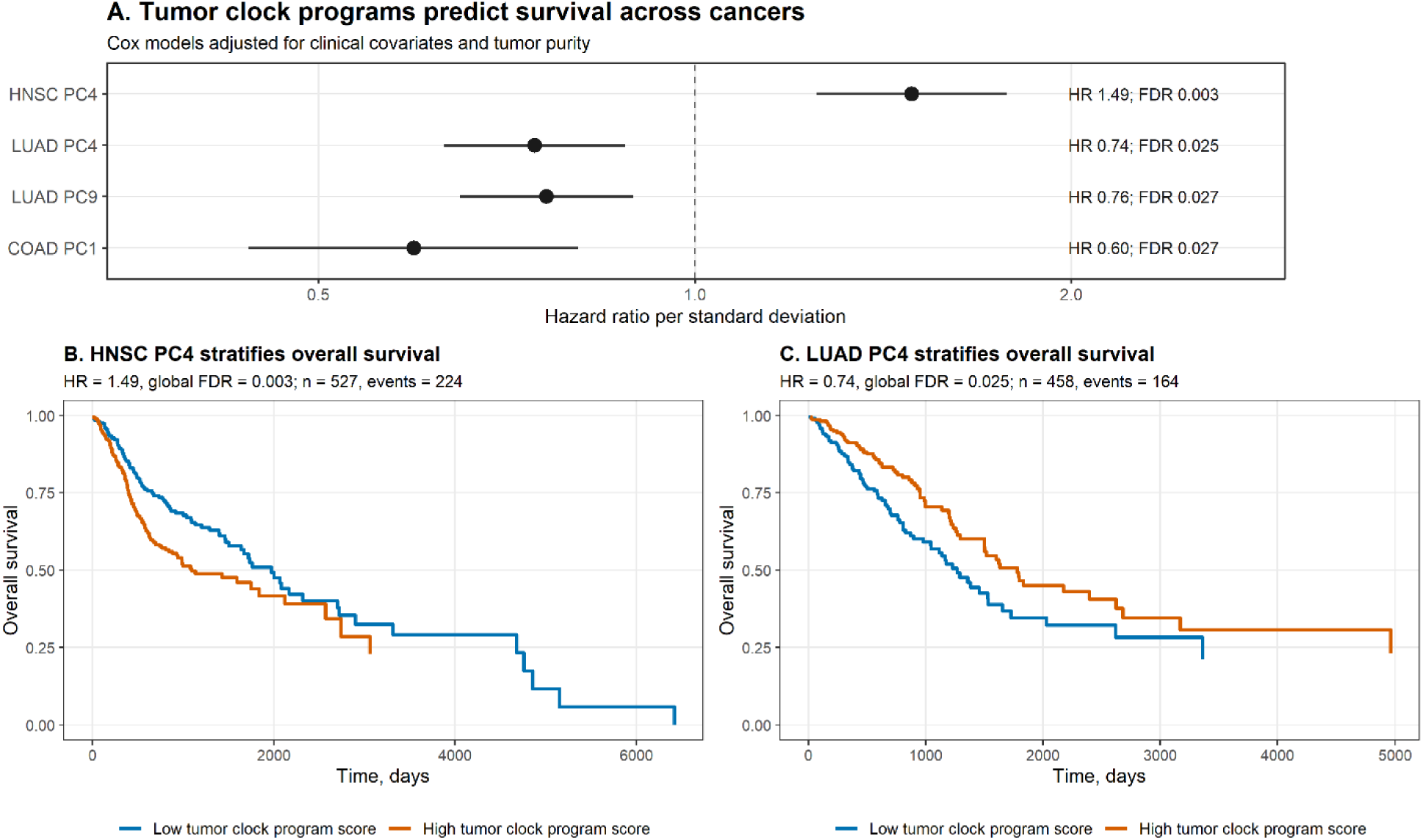
Tumor methylation programs predict survival across cancers. (A) Adjusted Cox models identified four tumor methylation programs associated with overall survival after global FDR correction: HNSC PC4 (HR 1.49; FDR=0.003), LUAD PC4 (HR 0.74; FDR=0.025), LUAD PC9 (HR 0.76; FDR=0.027), and COAD PC1 (HR 0.60; FDR=0.027). Hazard ratios are per one-standard-deviation increase in program score. (B,C) Representative Kaplan-Meier curves show worse survival with high HNSC PC4 and improved survival with high LUAD PC4; patients were split by median program score.

### Prognostic methylation programs encode cancer biology

GSVA analysis linked survival-associated tumor methylation programs to specific transcriptional signatures. Associations were adjusted for age, sex, stage, tumor purity, and proliferation/instability measures. The adverse HNSC PC4 program was positively associated with unfolded protein response and EMT activity (UPR: partial r=0.398, FDR=1.69×10^−20^; EMT: partial r=0.335, FDR=1.04×10^−14^). Protective programs showed inverse associations: LUAD PC4 was negatively associated with UPR and IFN-γ response (partial r=-0.242 and −0.155; FDR=8.58×10^−7^ and 0.002), and COAD PC1 was negatively associated with IFN-γ response, EMT, and stemness (partial r=−0.439, −0.180, and −0.129; all FDR≤0.034). LUAD PC9 had no significant adjusted GSVA associations.

**Fig. 5.**
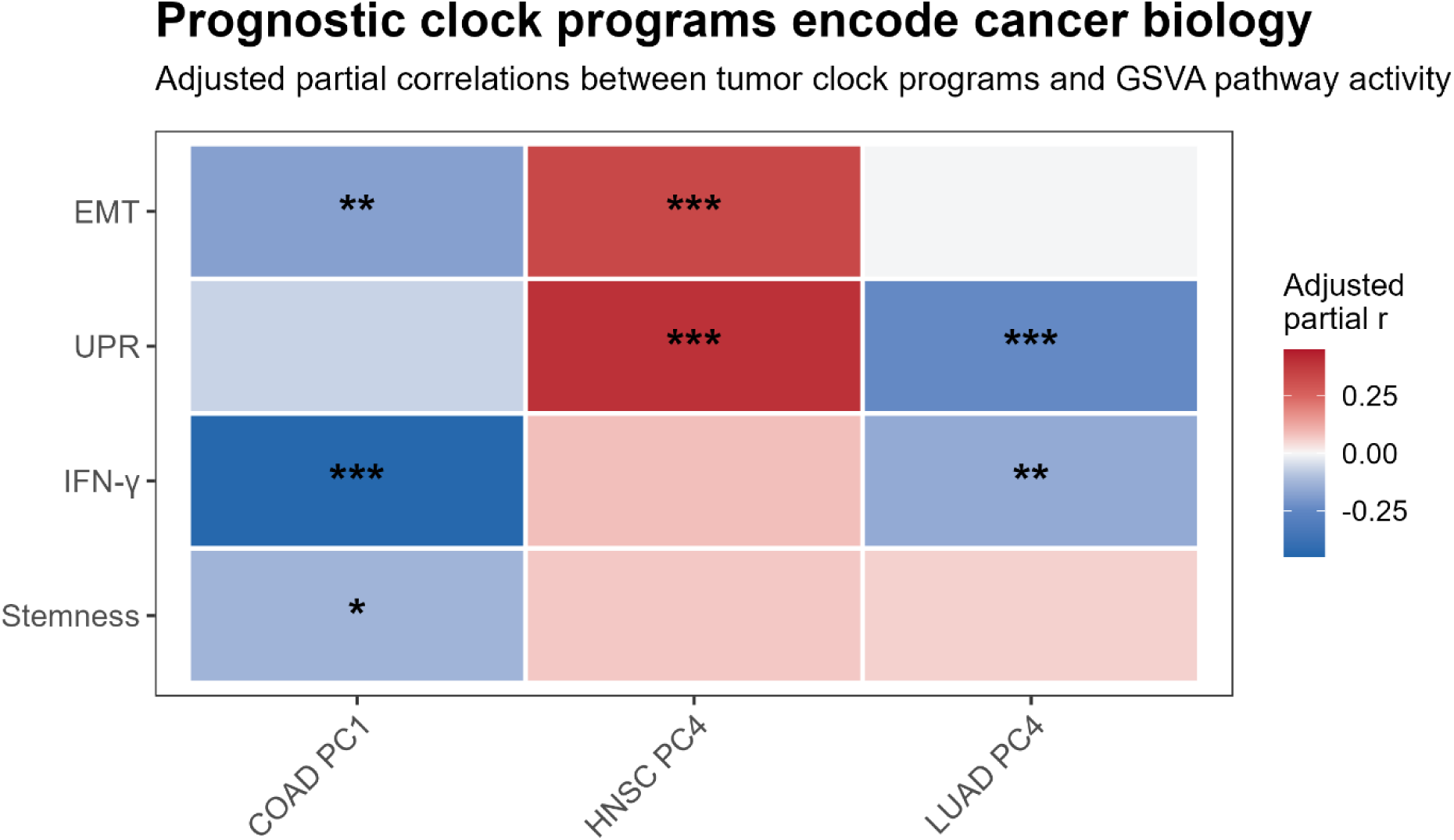
Prognostic methylation programs encode cancer biology. Heatmap showing associations between survival-associated tumor methylation programs and GSVA transcriptional signatures. Colors indicate adjusted partial correlation coefficients after adjustment for age, sex, stage, tumor purity, and proliferation/instability measures. Asterisks indicate FDR-significant associations after correction across signatures within each program. HNSC PC4, linked to poorer survival, was positively associated with unfolded protein response and EMT activity (UPR r=0.398, FDR=1.69×10^−20^; EMT r=0.335, FDR=1.04×10^−14^). COAD PC1 and LUAD PC4, linked to improved survival, showed inverse associations with IFN-γ response, EMT, stemness, and UPR (COAD PC1: IFN-γ r=™0.439, EMT r=™0.180, stemness r=™0.129; LUAD PC4: UPR r=™0.242, IFN-γ r=™0.155; all FDR≤0.034).

### Prognostic methylation programs persist after differentially methylated region control

Tumor-normal differentially methylated regions (DMRs) might explain prognostic tumor methylation programs by recapitulating generic cancer-associated methylation shifts. After controlling for DMR, the original tumor methylation programs persisted across all cancers.

PC1-PC10 explained 35.3%–47.2% of clock-CpG variance, including 43.6% in LUAD, 46.8% in HNSC, and 40.9% in COAD.

The original survival-linked programs also remained prognostic after controlling for DMR. In Cox models adjusted for age, sex, stage, tumor purity, and proliferation/instability measures, HNSC PC4 was associated with poorer survival (HR=1.40, FDR=7.78×10^−4^), while LUAD PC4, LUAD PC9, and COAD PC1 were associated with improved survival (HR=0.75, 0.81, and 0.67; FDR=0.001, 0.015, and 0.015, respectively).

**Fig. 6.**
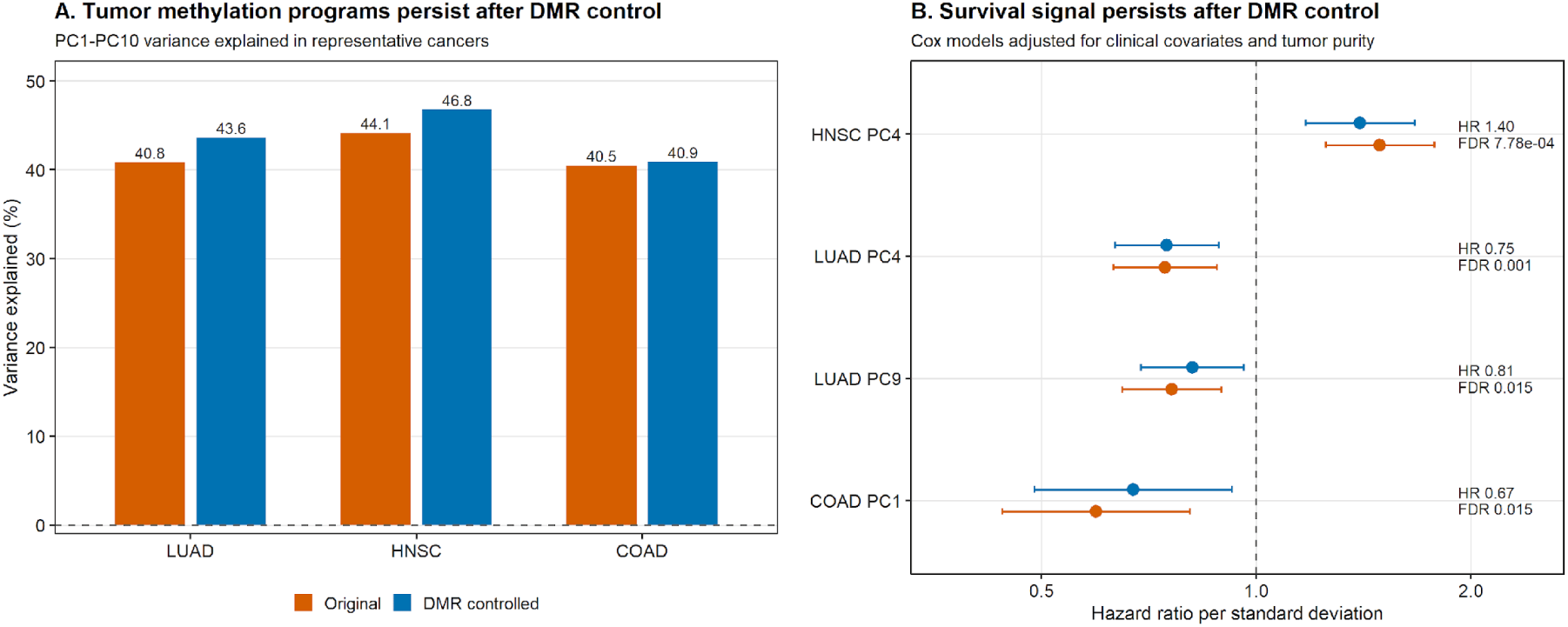
Prognostic tumor methylation programs persist after DMR control. (A) Tumor methylation programs captured 43.6% of clock-CpG variance in LUAD, 46.8% in HNSC, and 40.9% in COAD after DMR control. (B) Adjusted Cox models showed that the four survival-linked programs remained prognostic after DMR control: HNSC PC4 (HR 1.40; FDR=7.78×10^−4^), LUAD PC4 (HR 0.75; FDR=0.001), LUAD PC9 (HR 0.81; FDR=0.015), and COAD PC1 (HR 0.67; FDR=0.015). Hazard ratios are per one-standard-deviation increase in program score. FDR values in panel B were corrected across the four original survival-linked programs.

### External Validation

TCGA-BRCA tumor methylation programs were evaluated in GSE72308, an independent BRCA cohort, using the same TCGA-derived program definitions. Three TCGA-BRCA programs were nominally associated with survival after adjustment for age, sex, stage, tumor purity, proliferative history, and global methylation instability: PC2, PC4, and PC6. All three were associated with improved survival in TCGA-BRCA (PC2: HR 0.64, 95% CI 0.49–0.84, p=0.0015; PC4: HR 0.70, 95% CI 0.55–0.87, p=0.0019; PC6: HR 0.80, 95% CI 0.65–0.99, p=0.036). In GSE72308, PC4 and PC6 reproduced in the same protective direction (PC4: HR 0.68, 95% CI 0.53–0.87, p=0.0023; PC6: HR 0.72, 95% CI 0.54–0.97, p=0.030), while PC2 did not reproduce externally (HR 0.97, 95% CI 0.74–1.28, p=0.838).

**Fig. 7.**
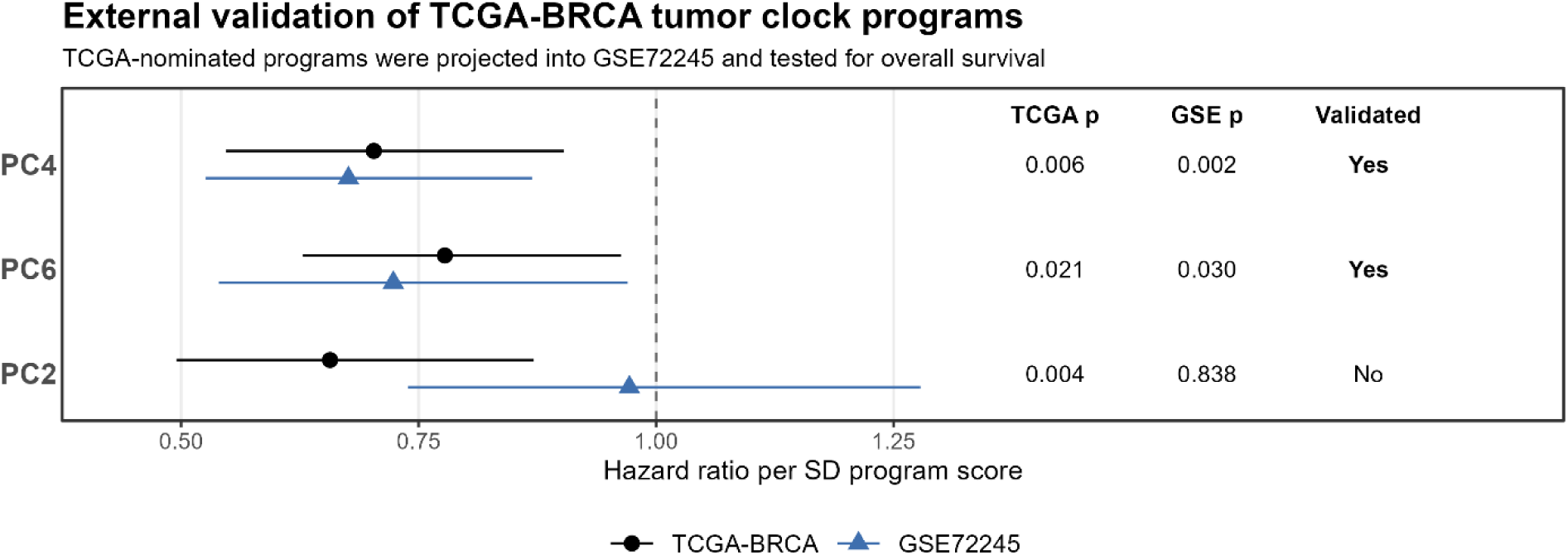
External validation of TCGA-BRCA tumor methylation programs. TCGA-BRCA programs nominally associated with overall survival after adjustment for age, sex, stage, tumor purity, proliferative history, and global methylation instability were evaluated in GSE72308 using the same TCGA-derived program definitions. Points show hazard ratios per one-standard-deviation increase in program score; horizontal lines show 95% confidence intervals. PC4 and PC6 reproduced in GSE72308 in the same protective direction, whereas PC2 did not reproduce externally.

## Discussion

Epigenetic clocks have traditionally been interpreted in cancer through methylation age or age acceleration [5-8]. This framing assumes that clock CpGs continue to behave as an aging system in tumors, such that their main cancer-relevant signal is whether a tumor appears epigenetically older or younger. Our results argue against that assumption. Tumors did not show a reproducible directional shift in Horvath age acceleration. Instead, they showed an order-of-magnitude increase in age-acceleration variance relative to normal tissues. More importantly, none of the five scalar clock measurements tested across nine cancers predicted survival even nominally after adjustment for age, sex, and stage. In tumors, the clock no longer behaves like an age measurement. Its cancer-relevant information emerges only when clock CpGs are analyzed as a disrupted methylation system.

This raises the central question of what clock CpGs represent once the aging axis breaks. Clock CpGs may reflect several different processes in normal tissue: causal aging mechanisms, downstream consequences of systemic aging, stochastic methylation drift, or regulatory states linked to development and cell identity [8, 17-18]. Cancer, however, acts on all of these at once [9-11]. Tumors accumulate methylation disorder, reactivate developmental programs, adopt stem-like states, and transition between adaptive cell states. Under this model, the informative signal is not a uniform shift toward older or younger methylation age, but a reorganization of relationships among clock CpGs. Analyzing those relationships can therefore reveal prognostic and tumor-state information that scalar clock analyses discard.

We therefore stopped treating Horvath clock CpGs as a fixed age predictor and analyzed them as a coordinated methylation system. PCA identified the major patterns of Horvath CpG covariation within each cancer type, which we define as tumor methylation programs. We then assigned each sample a tumor methylation program score, reflecting how strongly that sample exhibits the corresponding coordinated methylation pattern. Across nine cancers, the ten programs analyzed captured approximately one-third to one-half of clock-CpG variance, and this structure remained after accounting for tumor purity, proliferative history, and global methylation instability. Thus, the clock-CpG signal is not independent CpG-level noise or a trivial reflection of major tumor-level confounders.

These programs succeeded where scalar clocks failed. After full adjustment for clinical variables, tumor purity, proliferative history, and global methylation instability, tumor methylation program scores produced 15 nominal survival associations, compared with zero nominal associations for scalar clock measurements after clinical adjustment. Four of these associations remained significant after FDR correction across programs and cancers: HNSC PC4 was associated with poorer survival, whereas LUAD PC4, LUAD PC9, and COAD PC1 were associated with improved survival. In separate clinically adjusted models, proliferative history and global methylation instability produced no nominal survival associations across 17 tests, whereas tumor methylation programs produced 15 nominal associations and four FDR-significant associations despite full adjustment for both measures. This benchmark is meaningful because proliferative activity and global methylation instability are not arbitrary covariates. Proliferation is a clinically used marker of tumor aggressiveness, and global methylation instability has been linked to genomic instability, tumor aggressiveness, and clinical outcomes [19-21]. External validation showed that the TCGA-nominated survival programs were not dataset-specific correlations. Two of three TCGA-BRCA survival associations reproduced in GSE72308 in the same protective direction.

Transcriptional analysis supported the same interpretation. Survival-associated tumor methylation programs were linked to specific GSVA signatures after adjustment for age, sex, stage, tumor purity, and proliferative history. The adverse HNSC PC4 program was positively associated with UPR and EMT, whereas protective LUAD PC4 and COAD PC1 showed inverse associations with selected pathway signatures, including UPR, IFN-γ response, EMT, and stemness. The directions were coherent: adverse programs correlated positively with aggressive pathway activity, whereas protective programs correlated negatively. These pathway associations link the prognostic methylation programs to recognizable tumor states, rather than to methylation variation alone.

Tumor methylation programs also differ from conventional differentially methylated region (DMR) biomarkers. DMR approaches have been central to cancer epigenetics because many tumors show reproducible regional hypermethylation or hypomethylation, such as the CpG Island Methylator Phenotype (CIMP) in colorectal cancer [22]. But DMRs capture the most visible first-order methylation change: loci that shift up or down between biological states. CpGs may also carry higher-order information through coordinated relationships across loci, much as genes form expression programs beyond single-gene differential expression. A key concern was that tumor methylation programs might simply be recovering this lower-order DMR signal rather than genuine CpG covariation. To rule this out, we removed tumor-normal mean methylation at each individual Horvath CpG and evaluated whether the original programs still captured substantial variance and remained prognostic. The same programs continued to explain comparable clock-CpG variation and retained their survival associations.

The major limitation of this study is that PCA is a blunt instrument for learning tumor methylation programs. PCA identifies linear patterns that explain large sources of variation, but the most prognostic or biologically informative CpG relationships may not be the largest sources of variation. More targeted approaches such as supervised latent-variable models or pathway-informed methylation representations may identify stronger and more interpretable tumor programs. This study should be interpreted as a proof of concept. Even a simple unsupervised decomposition of Horvath clock CpGs reveals tumor methylation structure with survival and transcriptional relevance.

This study does not establish that Horvath clock CpGs are the optimal CpG set for tumor program analysis. Horvath CpGs were selected for age prediction, not cancer biology, and they are not linked by design to a single coherent biological function. Other biologically defined CpG sets may produce stronger tumor programs. For example, second generation epigenetic clocks such as PhenoAge and GrimAge, which are trained to incorporate mortality and indicators of health in order to predict lifespan and health rather than chronological age, may reveal tumor programs causally linked to prognosis and survival [23-25]. The advantage of Horvath CpGs here is that they provide a well-characterized methylation feature set with a strong normal-tissue baseline. Future work should compare clock CpGs against other predefined CpG sets and matched background CpG sets.

In conclusion, this study argues that epigenetic clocks in cancer should not be interpreted as measures of whether tumors are biologically older or younger. In tumors, Horvath clock CpGs are better understood as a methylation system that can be reorganized into coordinated tumor programs. Analyzing that structure reveals prognostic and transcriptional value that scalar clock analyses discard.

## Data Availability

TCGA methylation data are available online at the UCSC Xena browser. TCGA patient clinical data are available online at the GDC Data Portal. The validation breast cancer cohort (GSE72308) data are available online on the Gene Expression Omnibus repository. All data produced in the present study are contained in the manuscript.

https://portal.gdc.cancer.gov/

https://xenabrowser.net/datapages/

https://www.ncbi.nlm.nih.gov/geo/query/acc.cgi?acc=GSE72308

## References

1. Locke WJ, Guanzon D, Ma C, Liew YJ, Duesing KR, Fung KY, Ross JP. DNA methylation cancer biomarkers: translation to the clinic. Frontiers in genetics. 2019 Nov 14;10:1150.

2. Saha D, Kanjilal P, Kaur M, Menon SV, Ashraf A, Kumar MR, Alqahtani T, Atteri S, Uti DE, Dhara B. Transforming cancer diagnostics: The emergence of liquid biopsy and epigenetic markers. MedComm. 2025 Sep;6(9):e70388.

3. Leygo C, Williams M, Jin HC, Chan MW, Chu WK, Grusch M, Cheng YY. DNA methylation as a noninvasive epigenetic biomarker for the detection of cancer. Disease markers. 2017;2017(1):3726595.

4. Liang R, Tang Q, Chen J, Zhu L. Epigenetic clocks: beyond biological age, using the past to predict the present and future. Aging and disease. 2024 Dec 13;16(6):3520–3545.

5. Beynon RA, Ingle SM, Langdon R, May M, Ness A, Martin RM, Suderman M, Ingarfield K, Marioni RE, McCartney DL, Waterboer T. Epigenetic biomarkers of ageing are predictive of mortality risk in a longitudinal clinical cohort of individuals diagnosed with oropharyngeal cancer. Clinical Epigenetics. 2022 Dec;14(1):1.

6. Fransquet PD, Wrigglesworth J, Woods RL, Ernst ME, Ryan J. The epigenetic clock as a predictor of disease and mortality risk: a systematic review and meta-analysis. Clinical epigenetics. 2019 Dec;11(1):62.

7. Hong C, Yang S, Wang Q, Zhang S, Wu W, Chen J, Zhong D, Li M, Li L, Li J, Yu H. Epigenetic age acceleration of stomach adenocarcinoma associated with tumor stemness features, immunoactivation, and favorable prognosis. Frontiers in genetics. 2021 Mar 18;12:563051.

8. Yu M, Hazelton WD, Luebeck GE, Grady WM. Epigenetic aging: more than just a clock when it comes to cancer. Cancer research. 2020 Feb 1;80(3):367–74.

9. Saini A, Gallo JM. Epigenetic instability may alter cell state transitions and anticancer drug resistance. PLOS Computational Biology. 2021 Aug 23;17(8):e1009307.

10. Costa PM, Sales SL, Pinheiro DP, Pontes LQ, Maranhão SS, Pessoa CD, Furtado GP, Furtado CL. Epigenetic reprogramming in cancer: From diagnosis to treatment. Frontiers in cell and developmental biology. 2023 Feb 14;11:1116805.

11. Xu T, Li HT, Wei J, Li M, Hsieh TC, Lu YT, Lakshminarasimhan R, Xu R, Hodara E, Morrison G, Gujar H. Epigenetic plasticity potentiates a rapid cyclical shift to and from an aggressive cancer phenotype. International journal of cancer. 2020 Jun 1;146(11):3065–76.

12. Horvath S. DNA methylation age of human tissues and cell types. Genome biology. 2013 Oct;14(10):3156.

13. Thrush KL, Higgins-Chen AT, Liu Z, Levine ME. R methylCIPHER: a methylation clock investigational package for hypothesis-driven evaluation & research. BioRxiv. 2022 Jul 16:2022–07.

14. Teschendorff AE. A comparison of epigenetic mitotic-like clocks for cancer risk prediction. Genome Medicine. 2020 Jun 24;12(1):56.

15. Liberzon A, Birger C, Thorvaldsdóttir H, Ghandi M, Mesirov JP, Tamayo P. The molecular signatures database hallmark gene set collection. Cell systems. 2015 Dec 23;1(6):417–25.

16. Malta TM, Sokolov A, Gentles AJ, Burzykowski T, Poisson L, Weinstein JN, Kamińska B, Huelsken J, Omberg L, Gevaert O, Colaprico A. Machine learning identifies stemness features associated with oncogenic dedifferentiation. Cell. 2018 Apr 5;173(2):338–54.

17. Slieker RC, van Iterson M, Luijk R, Beekman M, Zhernakova DV, Moed MH, Mei H, Van Galen M, Deelen P, Bonder MJ, Zhernakova A. Age-related accrual of methylomic variability is linked to fundamental ageing mechanisms. Genome biology. 2016 Sep 22;17(1):191.

18. Bertucci-Richter EM, Shealy EP, Parrott BB. Epigenetic drift underlies epigenetic clock signals, but displays distinct responses to lifespan interventions, development, and cellular dedifferentiation. Aging (Albany NY). 2024 Jan 26;16(2):1002.

19. Kasprzak A. Prognostic biomarkers of cell proliferation in colorectal cancer (CRC): from immunohistochemistry to molecular biology techniques. Cancers. 2023 Sep 15;15(18):4570.

20. Salahi-Niri A, Zarand P, Mansouri N, Rastgou P, Yazdani O, Esbati R, Shojaeian F, Jahanbin B, Mohsenifar Z, Aghdaei HA, Ardalan FA. Potential of proliferative markers in pancreatic cancer management: A systematic review. Health Science Reports. 2025 Mar;8(3):e70412.

21. Bukovec D, Gjorgjioski B, Misheva MS, Kungulovski G. DNA methylation variability defines a fundamental dimension of tumor epigenomes linked to genomic instability, tumor aggressiveness, and clinical outcomes. bioRxiv. 2026 Mar 14:2026–03.

22. Worthley DL, Leggett BA. Colorectal cancer: molecular features and clinical opportunities. The Clinical Biochemist Reviews. 2010 May;31(2):31.

23. Crimmins EM, Klopack ET, Kim JK. Generations of epigenetic clocks and their links to socioeconomic status in the Health and Retirement Study. Epigenomics. 2024 Jul 17;16(14):1031–42.

24. Levine ME, Lu AT, Quach A, Chen BH, Assimes TL, Bandinelli S, Hou L, Baccarelli AA, Stewart JD, Li Y, Whitsel EA. An epigenetic biomarker of aging for lifespan and healthspan. Aging (albany NY). 2018 Apr 17;10(4):573.

25. Lu AT, Quach A, Wilson JG, Reiner AP, Aviv A, Raj K, Hou L, Baccarelli AA, Li Y, Stewart JD, Whitsel EA. DNA methylation GrimAge strongly predicts lifespan and healthspan. Aging (albany NY). 2019 Jan 21;11(2):303.

